# Multi-omic signatures of genetic mechanisms inform on type 2 diabetes biology and patient heterogeneity

**DOI:** 10.64898/2026.04.17.26351136

**Authors:** Magdalena Sevilla-González, Alan Magno Martínez-Muñoz, Paul A. Hanson, Sarah Hsu, Xingyang Wang, Kirk Smith, Zsu-Zsu Chen, Lukasz Szczerbinski, Varinderpal Kaur, Kent D. Taylor, Alexis C. Wood, Michael Y. Mi, Hui Li, Clemens Wittenbecher, Robert E. Gerszten, Stephen S. Rich, Jerome I. Rotter, Jun Li, Josep M. Mercader, Alisa K. Manning, Ravi V. Shah, Miriam S. Udler

**Affiliations:** Clinical and Translational Epidemiology Unit. Mongan Institute. Massachusetts General Hospital. Boston, MA, 02114, USA; Department of Medicine, Harvard Medical School, Boston, MA, 02115, USA; Programs in Metabolism and Medical & Population Genetics, The Broad Institute of MIT and Harvard, Cambridge, MA, 02142, USA; Diabetes Unit and Center for Genomic Medicine, Massachusetts General Hospital, Boston, MA, 02114, USA; Department of Epidemiology. Harvard T.H. Chan School of Public Health, Boston, MA, 02115, USA; Department of Preventive Medicine. Brigham & Women’s Hospital, Boston, MA, 02115, USA; Division of Cardiovascular Medicine, Beth Israel Deaconess Medical Center, Boston, MA, 02215, USA; The Institute for Translational Genomics and Population Sciences, Department of Pediatrics, The Lundquist Institute for Biomedical Innovation at Harbor-UCLA Medical Center, Torrance, CA 90502, USA; USDA/ARS Children’s Nutrition Research Center, Baylor College of Medicine-Department of Pediatrics, Houston, Texas, USA; Department of Life Sciences, Food and Nutrition Science, SciLifeLab, Chalmers University of Technology, Gothenburg, Sweden; Department of Genome Sciences, University of Virginia School of Medicine, Charlottesville, VA, 22908, USA; Division of Cardiology, Vanderbilt University Medical Center, Nashville, TN 37232, United States

## Abstract

Type 2 diabetes (T2D) is a heterogeneous disease shaped by genetic pathways related to insulin resistance and β-cell dysfunction, but how this heterogeneity is reflected molecularly remains unclear. We integrated partitioned polygenic scores (pPS) with proteomic and metabolomic profiling to define molecular signatures of T2D and their clinical relevance.

We analyzed UK Biobank participants with genomic, proteomic, and metabolomic data. In a disease-free training subset, we used LASSO regression to identify multi-omic signatures associated with each pPS by jointly modeling proteins and metabolites. In an independent testing set, we constructed multi-omic scores and examined their associations with clinical traits and diabetes-related outcomes. Mediation analyses were used to investigate putative causal pathways. Key findings were evaluated in the Multi-Ethnic Study of Atherosclerosis (MESA).

We identified distinct multi-omic signatures that capture the molecular architecture of T2D genetic risk across physiological subtypes. Compared with genetic scores alone, multi-omic pPS showed larger effect sizes and better disease discrimination. These scores recapitulated subtype-specific physiology and were associated with T2D risk. The Beta-Cell 2 multi-omic score showed marked stratification for insulin use, which was replicated in MESA, where it also predicted future insulin use. Mediation analyses implicated lipoprotein remodeling and fatty acid metabolism in the Lipodystrophy 1 cluster, accounting for 30–45% of the total effect of pPS on T2D risk.

Integrating process-specific genetic risk with circulating multi-omic profiles reveals biologically distinct endotypes of T2D and supports a framework for improved patient stratification and risk assessment.

## INTRODUCTION

Type 2 diabetes (T2D) affects more than 500 million individuals worldwide^1^ with profound consequences on longevity, quality of life, and morbidity. T2D exhibits clinical-biological heterogeneity, with substantial variability in onset, progression, and complications. At a physiologic level, heterogeneity relates to the balance between impaired insulin action and β-cell dysfunction.^2^ Genetic studies have recently attempted to address this heterogeneity through reproducible physiological polygenic subtypes of T2D that partition inherited risk into distinct but interrelated disease processes.^3–6^ While these polygenic instruments may enhance precision in subtyping, by construction, they consist of only genetic markers, and not the molecular biomarkers through which these genetically defined subtypes may manifest. Large-scale proteomic and metabolic profiling studies serve as dynamic, proximal readouts of metabolic and inflammatory processes,^7^ ^8^ ^9^ ^10^ ^11,12^, however, most studies have been conducted independent of underlying genetic architecture, limiting their ability to inform potential mechanisms of heterogeneity.

Integrating physiological polygenic pathway scores with circulating molecular profiles provides a principled framework for linking genetic risk to downstream biological pathways. Here, we integrated partitioned polygenic scores (pPS) with large-scale plasma proteomic and metabolomic profiling in a population-based cohort. We define multi-omic signatures of T2D genetic subtypes, evaluate their associations with metabolic traits and disease progression, and prioritize putative causal mechanisms. Our goal was to identify molecular signatures informed by distinct genetic subtypes of T2D (e.g. insulin resistance- and β-cell dysfunction-driven endotypes), linking these signatures to clinical and metabolic comorbidity, including progression to overt T2D, insulin requirement, and diabetes-related complications.

## RESULTS

### Study Population Characteristics

After applying all exclusion criteria to remove individuals with major chronic cardiometabolic diseases and cancer, 38,205 participants in the UK Biobank (UKBB) with paired metabolomic and proteomic data were included in the analytic cohort. Baseline characteristics of the training and testing cohort are presented in **Supplementary Table 1**.

### Multi-omic signatures of T2D genetic subtypes

LASSO regressions across the 12 pPS and the global T2D PRS in the UKBB training set identified distinct multi-omic signatures that delineate the molecular architecture of T2D genetic risk and its physiological subtypes (**Supplementary Fig. 1A; Supplementary Tables 2–3**). All models remained significant under permutation testing, indicating that predictive performance was unlikely to arise by chance (all permutation P < 1 × 10□□).

Partitioned polygenic scores (pPS) delineated distinct circulating proteomic and metabolomic profiles. Clusters related to insulin production and processing, including Beta Cell 1 and Beta Cell 2, showed concordant associations with regulators of β-cell function and lipid metabolism. Both were associated with lower levels of CEACAM1, a key mediator of insulin clearance ^13^, and a higher omega-6 to omega-3 ratio, indicating lipid remodeling and β-cell stress that may impair insulin secretory capacity (**Figure 1**). On the other hand, insulin resistance–related clusters exhibited distinct but complementary molecular profiles. Both Lipodystrophy 1 and Lipodystrophy 2, which reflect metabolically deleterious fat distribution, were associated with lower circulating levels of the adipokine leptin (LEP), consistent with a lipodystrophy-like phenotype. ^14^ Yet, their metabolomic signatures diverged: Lipodystrophy 2 was enriched in polyunsaturated fatty acids (e.g., DHA, LA), while Lipodystrophy 1 showed higher omega-6 fatty acids, suggesting differential lipid signaling. Additionally, ARG1, a regulator of nitric oxide metabolism and inflammation^15^, showed opposing associations across clusters—positive in Lipodystrophy 2 but negative in Lipodystrophy 1—highlighting differences in inflammatory tone and vascular metabolism. Consistent with these observations, Lipodystrophy 1 positively weighted proteins were enriched for interleukin signaling, whereas Lipodystrophy 2 negatively weighted proteins were enriched for cytokine and TNF receptor signaling (**Supplementary Figure 3**). The multi-omic signature of the obesity cluster was characterized by higher LEP (in contrast to the Lipodystrophy clusters) and lower PLA2G7, a phospholipase linked to macrophage activation and vascular inflammation ^16^, consistent with the chronic low-grade inflammation accompanying excess adiposity. Finally, the Liver Lipid cluster showed a profile of disrupted lipid turnover and hepatic metabolic stress, marked by elevated saturated fatty acids and proteins such as CGREF1, IGFBP2, and SMPD1, implicating pathways in lipid signaling^17^, glucose^18^, and sphingolipid pathways (**Figure 1**). Enrichment in interleukin signaling pointed to an inflammatory component, while lower linoleic acid suggested reduced unsaturated lipid availability (**Supplementary Figure 3**). Collectively, these results supported distinct molecular architecture underlying pPS genetic liability.

**Figure 1.**
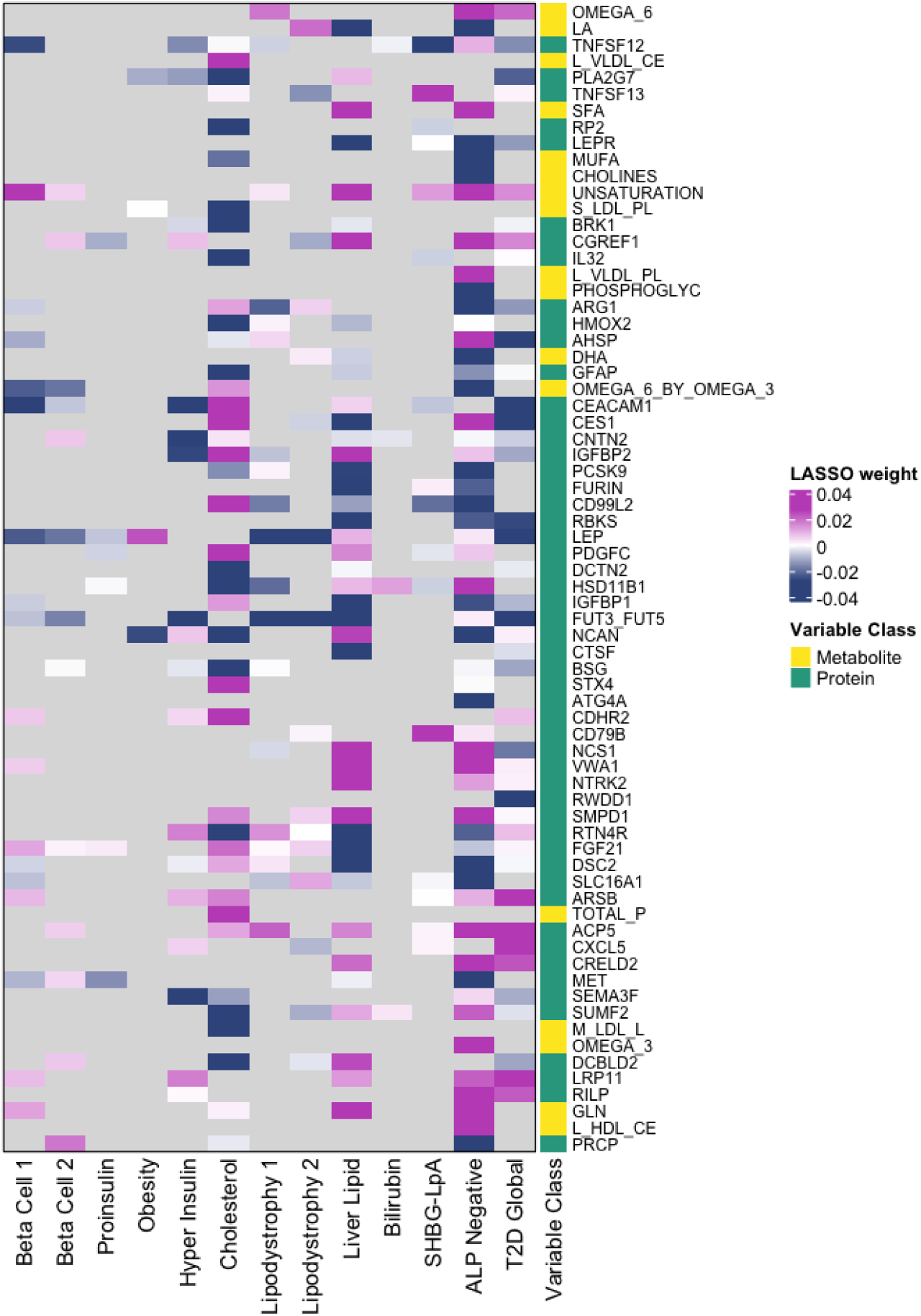
Top weighted metabolites and proteins contributing to multi-omic pPS scores across T2D subtypes in UKBB. Heatmap showing the top 50 molecules with the largest absolute LASSO coefficients contributing to each partitioned polygenic score (pPS). Columns represent the 12 T2D genetic subtypes and the global T2D score, and rows represent individual metabolites (yellow) and proteins (teal). Colors reflect standardized LASSO weights, where positive coefficients (pink) indicate higher molecule abundance associated with higher subtype score, and negative coefficients (navy) indicate inverse relationships. Gray color represents no presence of the value across the pPS.

To assess the extent to which pathway-specific genetic clusters capture molecular features shared with overall T2D risk, we evaluated overlap between each pPS-derived signature and the global T2D feature set. Most genetic clusters showed significant enrichment for metabolite and protein features included in the global signature (**Supplementary Fig. 1B; Supplementary Table 4**). The strongest enrichment was observed for the Hyperinsulin and Beta-Cell 1 clusters, followed by clusters related to liver–lipid metabolism and lipodystrophy (all FDR-adjusted P < 0.05), indicating substantial shared molecular features implicated across key T2D-related mechanisms.

### Multi-omic Scores Provide Complementary Information Beyond Genetic Risk

To leverage identified molecular signatures for clinical prediction, we constructed multi-omic pPS scores by aggregating weighted metabolites and proteins selected for each genetic subtype. Pairwise correlations between genetic and multi-omic scores were modest (**Figure 2A**), suggesting that the multi-omic scores capture information that is only partially shared with genetic risk. In mutually adjusted models that included both the genetic pPS and its corresponding multi-omic score, the multi-omic signatures retained independent associations with T2D and had larger effect sizes (**Figure 2B**), indicating that molecular abundances capture additional, disease-relevant variation beyond inherited genetic risk. For all pPS domains, models including multi-omic scores significantly enhanced discrimination over the base model (**Figure 2C**). Compared with the corresponding genetic scores, multi-omic scores showed stronger associations with subtype-specific clinical endotypes, with larger effect sizes in both positive and negative directions in individuals without T2D (**Figure 2D**), and those with T2D (**Supplementary Figure 3**). No significant interactions between the genetic pPS and the multi-omic scores were observed.

**Figure 2.**
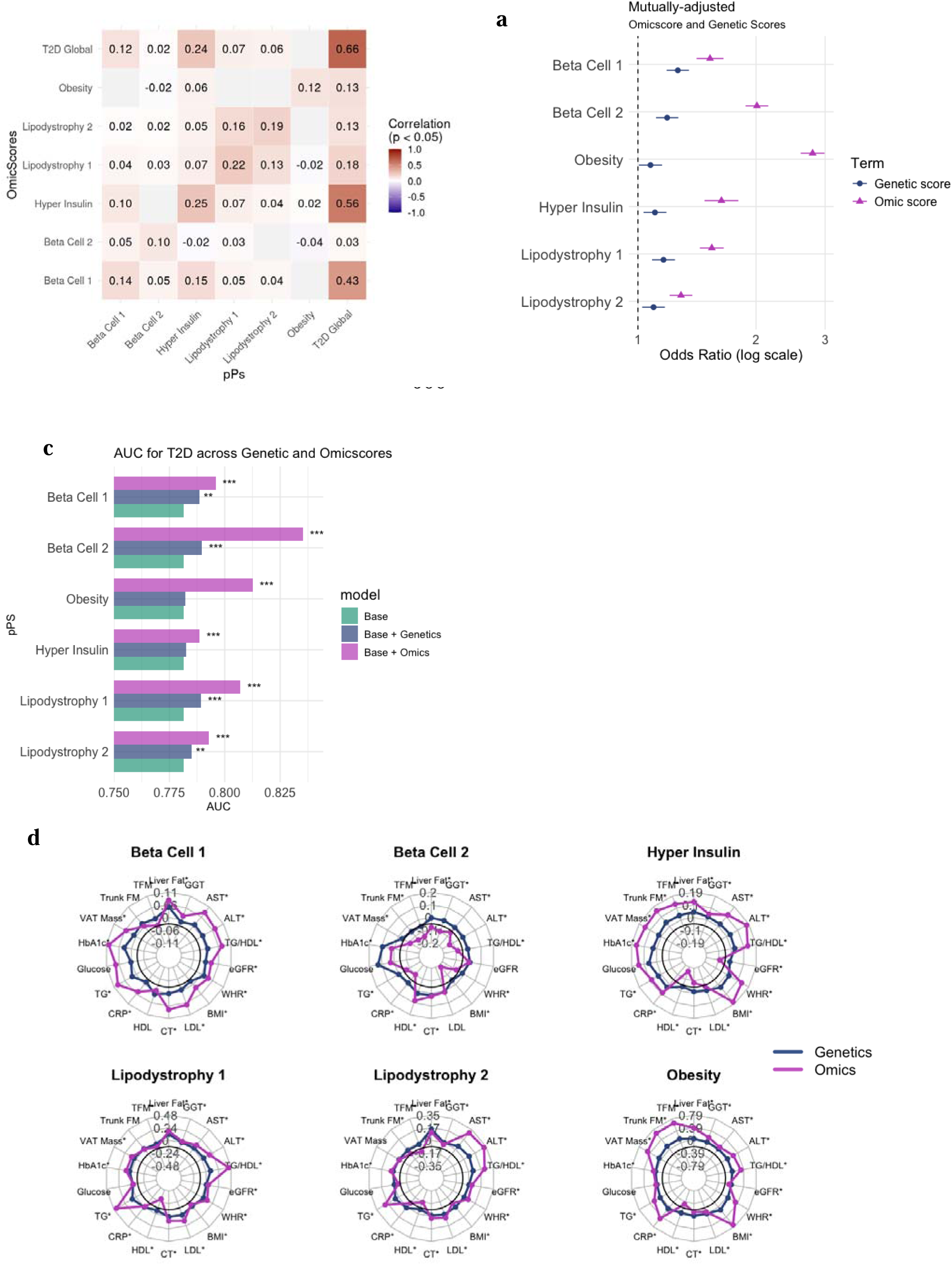
Comparison of genetic and multi-omic scores across T2D subtype-related pathways. (**A**) Spearman correlation matrix comparing partitioned polygenic scores (pPS) with corresponding multi-omic pPSs. Plot displays only *p<0.05* correlations. (**B**) Mutually adjusted logistic regression models showing associations of genetic and multi-omic scores with prevalent T2D. Models were adjusted for age, sex, 10 genetic principal components. Type 2 Diabetes cases N=1004. Error bars represent 95% confidence intervals. (**C**) Area under the curve (AUC) comparison for T2D classification models, including: (i) a base model (age, sex, BMI, and 10 principal components), (ii) base + genetic pPS, and (iii) base + multi-omic pPS. Statistical significance (DeLong test) reflects improvement in model discrimination relative to the base model (***P<0.001; **P<0.01). (**D**) Radar plots show standardized β coefficients for the associations of pathway-specific genetic scores (blue) and corresponding multi-omic scores (magenta) with inverse-normalized clinical traits in individuals without type 2 diabetes (N=15,527). The bold black line denotes the null effect (β = 0). Each panel corresponds to one T2D subtype pathway: Beta Cell 1, Beta Cell 2, Hyper Insulin, Lipodystrophy 1, Lipodystrophy 2, and Obesity. Effect estimates are shown per 1-SD higher score. Asterisks indicate associations significant after Bonferroni correction for multiple comparisons. TFM: Total Fat Mass, Trunk FM: Trunk Fat Mass, VAT: Visceral Adiposity, TG: Triglycerides, CRP: C-Reactive Protein, CT: Total Cholesterol, WHI: Waist-hip-ratio. Models were adjusted for age, sex, and the first ten genetic principal components and fasting hours. Associations with random glucose were restricted to participants with >6 hours of fasting.

### Multi-omic Scores Capture Endotypes Across Metabolic Disease Progression

To determine whether multi-omic pPS profiles change in relation to disease progression, we examined the distribution of each score across cross-sectional metabolic health stages (**Figure 3**). All multi-omic scores showed clear, graded increases from normoglycemia to prediabetes and T2D (P < 0.001 for trend for all scores), indicating that the molecular signatures underlying each subtype track with worsening metabolic health.

**Figure 3.**
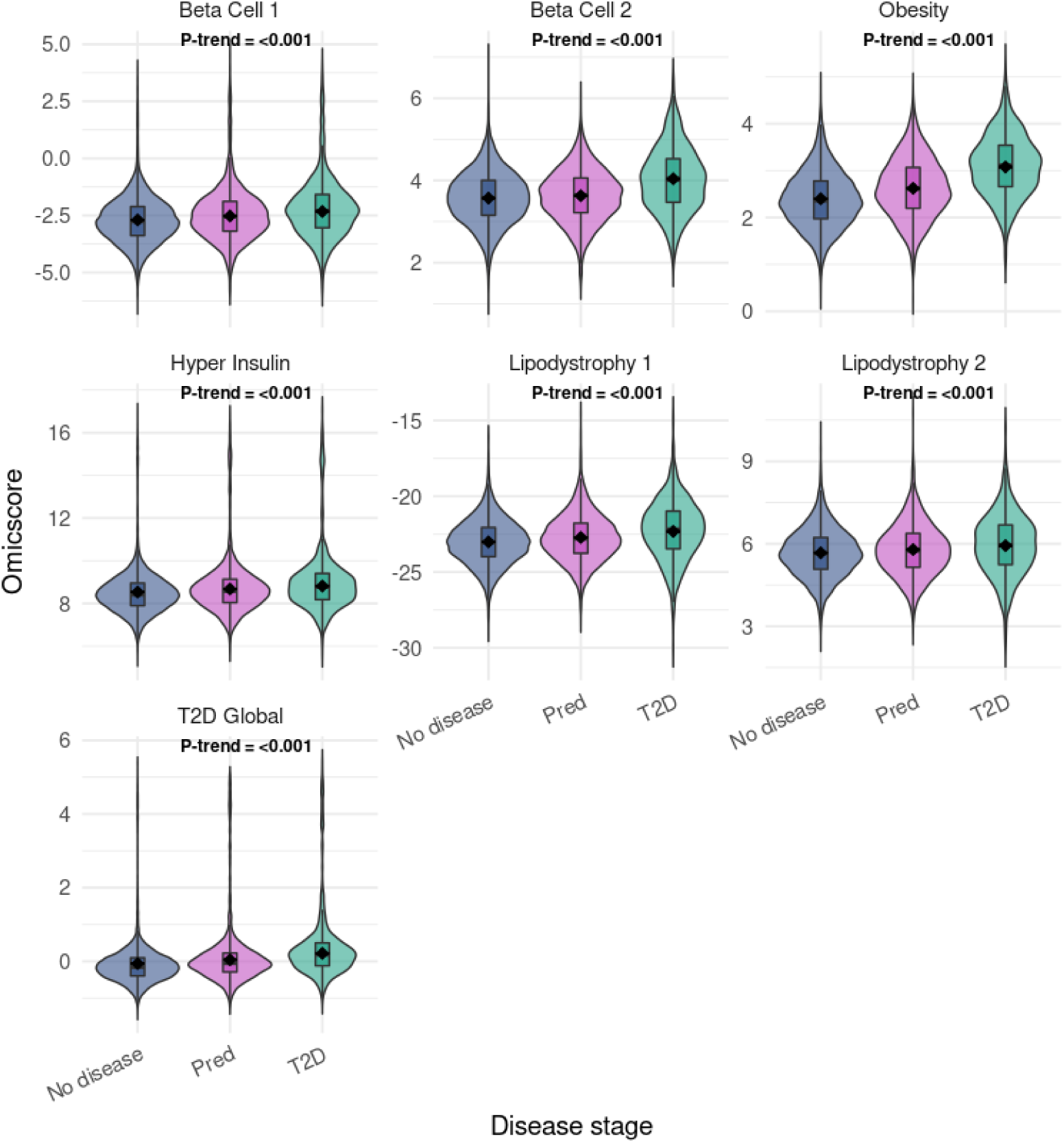
Multi-omic pPS scores across Disease Stages. Violin plots illustrating the distribution of pPS omics scores across three disease stages: ’No disease’ (N=15,527), ’Pred’ (prediabetes, N=3,579), and ’T2D’ (Type 2 Diabetes, N=1004). The plotted values represent Estimated Marginal Means (EMMs), which have been statistically adjusted for age, sex, and the first 10 genetic principal components (PCs) to account for confounding factors and population structure. The black dots within each violin indicate the median, and the thick black bars represent the interquartile range. P-trend values, indicating the significance of a linear trend across disease stages, are displayed for each omic score. A P-trend Bonferroni adjusted is considered statistically significant.

Furthermore, multi-omic pPS showed coherent associations with clinical traits that largely mirrored those of the genetic pPS (**Figure 4**). The Lipodystrophy 1 score was characterized by a higher waist-to-hip ratio, triglyceride-to-HDL (TG/HDL) ratio, liver fat, and visceral adiposity, together with lower total fat mass and BMI. The Lipodystrophy 2 score showed a similar insulin resistance-related pattern, with a higher TG/HDL ratio, in addition to liver fat and hepatic transaminases, lower total fat mass, and BMI. The Obesity score was associated with increased adiposity-related measures and a higher TG/HDL ratio. Notably, the insulin resistance profiles of the lipodystrophy and obesity clusters were replicated in MESA, where Lipodystrophy 1 and 2 were associated with higher HOMA-IR, and the Obesity cluster with higher BMI and HOMA-IR (**Supplementary Figure 4**).

**Figure 4.**
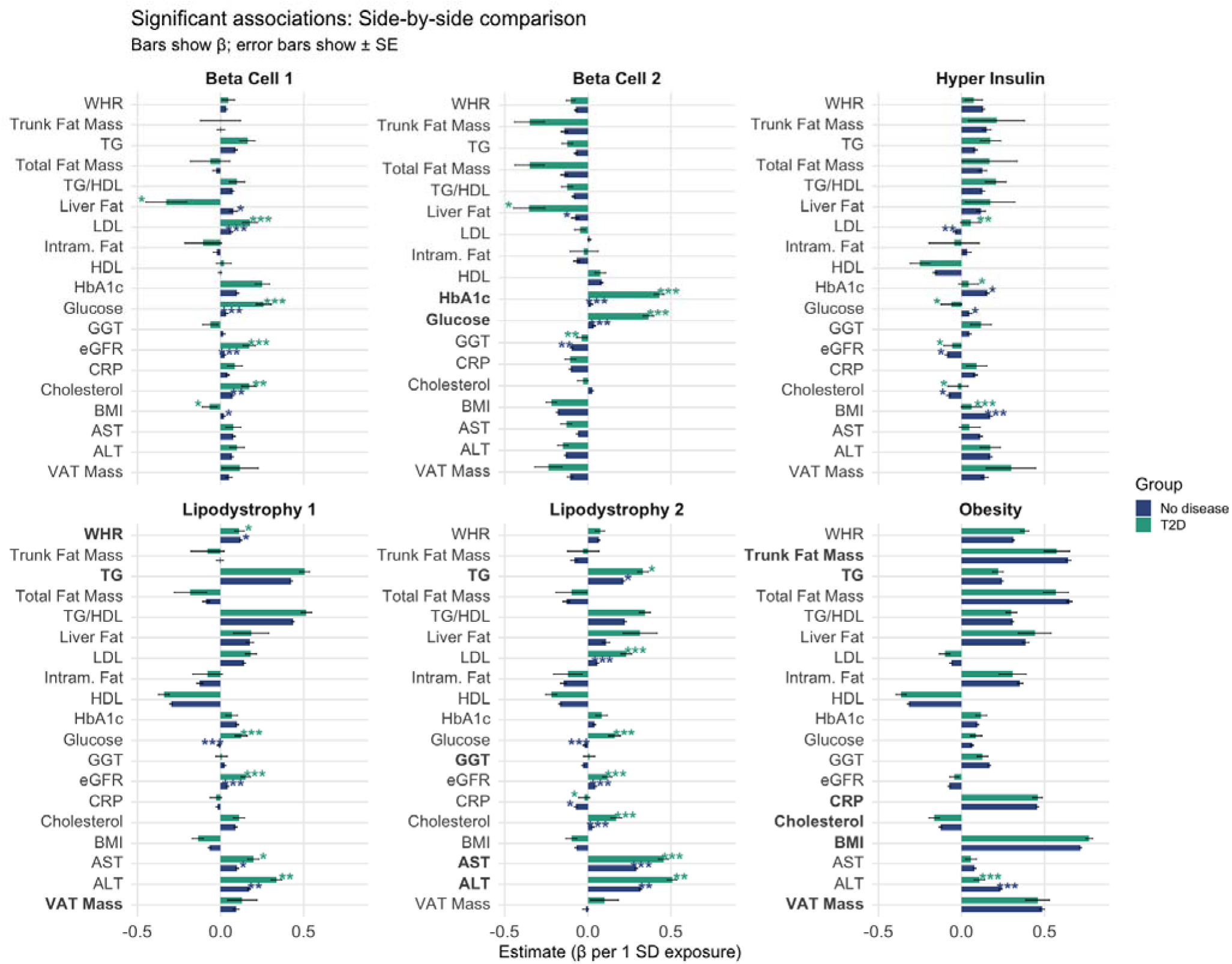
Multi-omic pPS scores across clinical traits stratified by diabetes status. Bar plots illustrate the associations between each multi-omic pPS and clinical traits in individuals with and without type 2 diabetes. Plot displays significant traits (Bonferroni-adjusted P < 0.05). Bars represent β-estimates from linear regression models adjusted for age, sex, the first ten genetic principal components and fasting hours; error bars indicate ± standard error. Associations with random glucose were restricted to participants with >6 hours of fasting. Trait labels shown in bold denote associations expected a priori based on known genetic pathways underlying each pPS. Asterisks indicate multi-omic pPS scores × disease stage interactions (No disease [N=15,527] vs. T2D [N=1,004]): * < 0.05; ** <0.01; *** <0.001.Clinical traits span metabolic and anthropometric domains, including inflammation (C-reactive protein [CRP]), adiposity (body mass index [BMI], waist circumference, hip circumference, waist-to-hip ratio), body composition (visceral adipose tissue, trunk fat mass, total fat mass, intramuscular fat, liver fat), lipid metabolism (triglyceride-to-HDL ratio, LDL-C, HDL-C, total cholesterol), kidney function (estimated glomerular filtration rate [eGFR]), and glycemic markers (fasting glucose and HbA1c). Blue bars indicate individuals without T2D, and green bars indicate individuals with T2D.

Stratified analyses revealed that several associations were amplified in individuals with T2D, with significant interaction effects indicating modification by disease status. In particular, β-cell–related clusters (Beta-Cell 1 and Beta-Cell 2) showed markedly stronger associations with glucose and HbA1c in the T2D subset, consistent with amplified β-cell dysfunction.

Hyperinsulin and Lipodystrophy 1 and 2 clusters exhibited stronger relationships with insulin resistance (TG:HDL), visceral and trunk adiposity, triglycerides, and inflammatory markers in individuals with T2D, reflecting more severe insulin-resistant phenotypes in the disease state. In contrast, the Obesity cluster showed stable associations with adiposity, fat distribution, insulin resistance (TG:HDL), and lipid traits across both T2D and non-T2D groups, suggesting an underlying metabolic architecture largely preserved with diabetes onset. Together, these findings indicate that multi-omic pPS capture physiologically meaningful endotypes whose clinical expression intensifies as individuals progress to overt T2D (**Figure 4**).

### Multi-omic scores are associated with T2D risk, and T2D-related complications

We next assessed whether multi-omic pPS were associated with T2D risk. Across all clusters, higher multi-omic scores were associated with greater T2D prevalence and outperformed the corresponding genetic pPS in capturing phenotypic risk, particularly at the highest deciles. In contrast to the linear gradient observed for genetic risk, multi-omic scores showed a pronounced threshold pattern, identifying a high-risk subgroup with up to threefold higher T2D prevalence than that defined by genetic scores alone, independent of BMI (**Supplementary Figure 5A**). This pattern was further supported in MESA, where protein-only and modified multi-omic scores showed broadly consistent associations with incident T2D (**Supplementary Figure 5B**).

Insulin use varied markedly across subtype-specific omic signatures in both continuous and top-decile analyses, with the strongest signal observed for the Beta-Cell 2 signature. Individuals in the highest Beta-Cell 2 multi-omic score decile had 14.69-fold higher odds of insulin use than the remainder of the cohort after adjustment for age, sex, and genetic ancestry (**Figure 5A**). This association remained after additional adjustment for diabetes duration, although with attenuation of effect size (OR, 2.17; 95% CI, 1.45–3.25) (**Supplementary Figure 6B**). In continuous analyses, each Standard Deviation (SD) increase in the Beta-Cell 2 multi-omic score was associated with 3.79-fold higher odds of insulin use (95% CI, 3.28–4.38) (**Supplementary Figure 6A**) in UKBB. A similarly strong association was observed for the corresponding proteomic score, whereas the genetic score alone showed only a modest effect. Obesity-related scores were also associated with insulin use, but with substantially smaller effect sizes, and these associations were no longer significant after adjustment for diabetes duration. Supportive findings were observed in MESA, where baseline protein-only and modified multi-omic scores were associated with insulin use cross-sectionally as well as with future insulin use at 10- and 15-year follow-up (**Figure 5B**), with effect sizes similar to those seen in UKBB (**Supplementary Figure 6A**).

**Figure 5.**
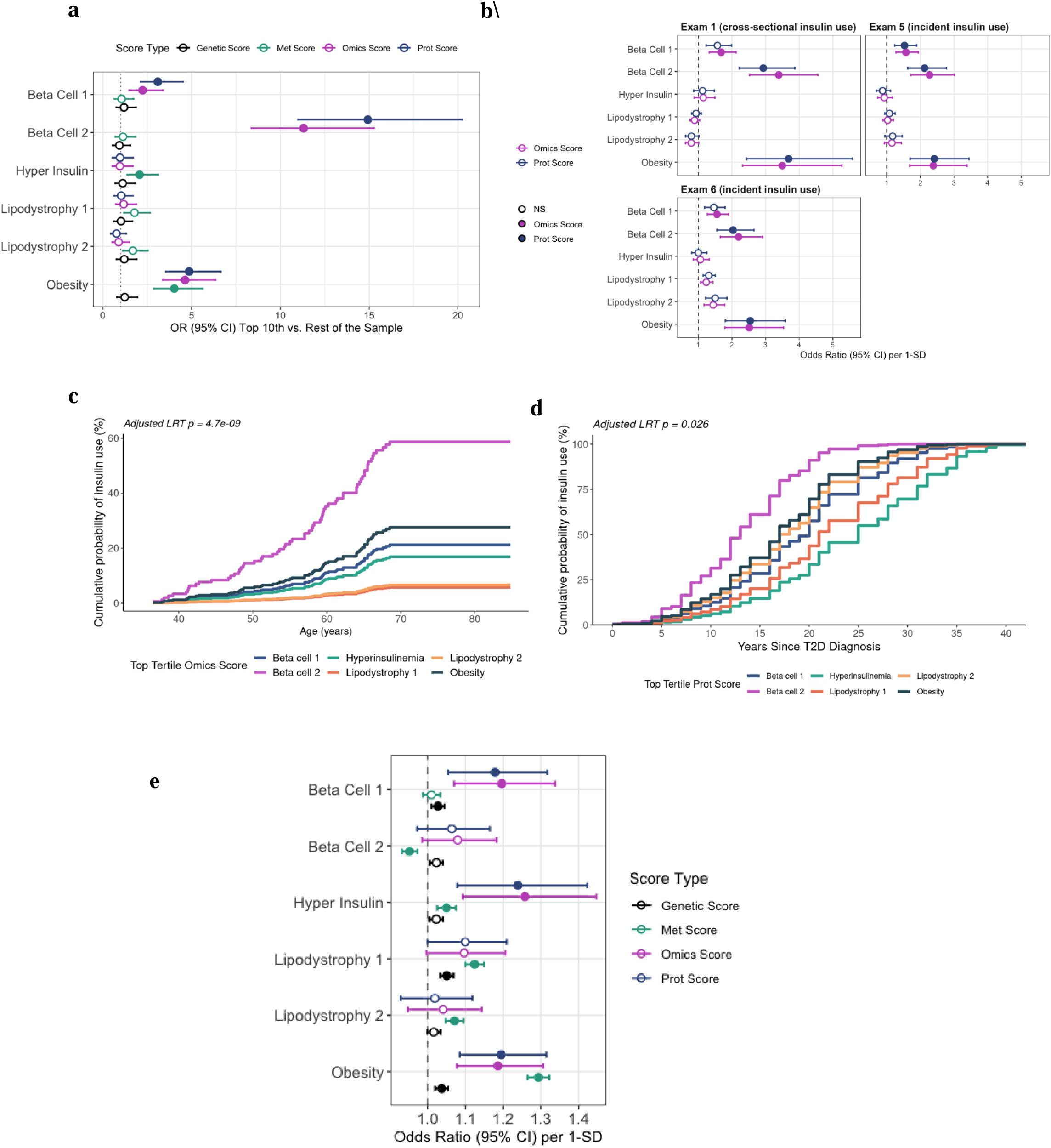
Associations between multi-omic pPS scores with T2D-related complications. **(A)** Forest plots display the associations of each genomic and multi-omic pPS with insulin use in UKBB when using the same sample size (Insulin cases: 180, Controls: 19,924). Estimates are shown as odds ratios (ORs) with 95% confidence intervals, adjusted for age, sex, and 10 principal components of ancestry. Effect sizes represent the contrast between individuals in the top 10th percentile of each score versus the remainder of the cohort. (**B**) Forest plots show odds ratios (ORs) and 95% confidence intervals for insulin use in relation to cluster-specific omics and proteomic scores at Exam 1 (baseline; cross-sectional), Exam 5 (∼10 years after baseline; new insulin cases only), and Exam 6 (∼15 years after baseline; new insulin cases only) in the full MESA cohort (n = 5,892). The number of insulin use cases was 105, 125, and 113 at Exams 1, 5, and 6, respectively. Filled points denote associations meeting the significance threshold (P < 0.001), whereas hollow points denote non-significant associations. All models were adjusted for age, sex, study site, and 10 principal components of ancestry. (**C**) Cumulative probability of insulin use by unique top-tertile multi-omic pPS subgroup among individuals with T2D in UKBB. Kaplan–Meier curves show the cumulative probability of insulin use according to unique top-tertile membership for each multi-omic pPS. Participants were assigned to mutually exclusive groups based on the subtype for which they were in the top tertile. The x-axis represents age in years, and the y-axis represents the cumulative probability of insulin use (%). Covariates include age, sex, and 10 principal components of ancestry and years living with T2D. Overall differences across groups were assessed using a global log-rank test. (**D**) Cumulative incidence curves for insulin use are shown according to subtype-specific omics scores across years since type 2 diabetes diagnosis in UKBB. Separation across curves was tested with the log-rank test, covariates include age, sex, genetic, and 10 genetic principal components. (**E**) Associations of pathway-specific genetic and molecular scores with incident cardiovascular disease (CVD) in UKBB. Forest plots show odds ratios (ORs) and 95% confidence intervals for associations between pathway-specific Genetic Score (cases = 14,535), MetScore (cases = 8,950), ProtScore (cases = 493), and Omics Score (cases = 487) and incident CVD. Points indicate effect estimates from logistic regression models, and horizontal lines denote 95% confidence intervals. Filled points highlight positive statistically significant associations (*p<0.004*). Models were adjusted for age, sex, statin use, smoking, and 10 genetic ancestry principal components.

We also assessed the cumulative probability of insulin use by age among individuals with T2D in UKBB and found marked separation in age at insulin initiation across unique top-tertile omics-pPS groups (global log-rank P = 7.2 × 10^-13^) (**Figure 5C**). The Beta-Cell 2 group showed by far the highest cumulative probability of insulin use, with 20% prescribed insulin by age 50 compared with less than 10% in all other groups. A similar pattern was seen with time from diabetes to diagnosis to insulin initiation, with the top tertile Beta-Cell 2 proteomic group again showing the earliest and steepest rise in cumulative insulin use and significant differences across cluster proteomic groups (L*RT P = 0.026*) (**Figure 5D**).

Furthermore, associations with incident CVD varied across subtype-specific scores and were generally stronger for molecular than for genetic scores. The strongest positive associations were observed for the Beta Cell 1, Hyper Insulin, and Obesity signatures, particularly for multi-omic scores (**Figure 5E**). Lipodystrophy 1 and 2 also showed significant positive associations for the metabolomic scores. These associations were largely unchanged after additional adjustment for T2D status (**Supplementary Figure 6C**).

### Causal mechanisms underlying multi-omic signatures and metabolic risk

To investigate how molecular intermediates translate genetic susceptibility into metabolic phenotypes, we conducted mediation analyses evaluating whether multi-omic features mediated associations between genetic pPS and regional adiposity traits or T2D (**Figure 6**).

**Figure 6.**
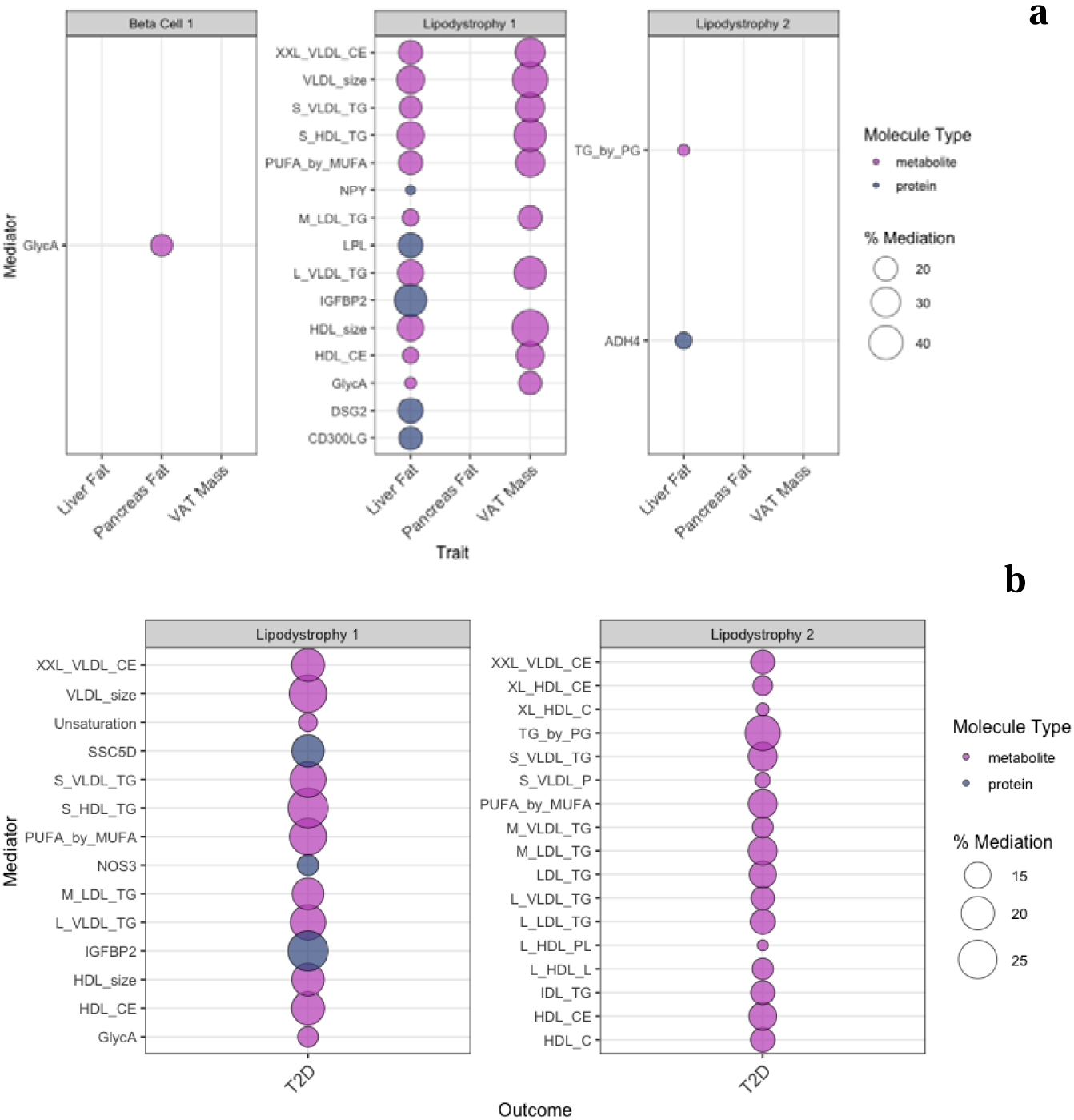
Mediation of genetic pPS effects on adiposity traits and T2D by multi-omic signatures. Bubble plots display statistically significant mediators linking partitioned polygenic scores (pPS) to (**A**) regional adiposity traits (liver fat, pancreatic fat, and VAT mass; Bonferroni-corrected *P* < 1×10□□) and (**B**) type 2 diabetes (T2D; Bonferroni-corrected *P* < 1×10□³). Each bubble represents a metabolite or protein included in the corresponding multi-omic pPS. The size of the bubble reflects the estimated proportion mediated, and color denotes molecular class (metabolite vs. protein). Only mediators meeting statistical significance after correction for multiple testing are shown. Results derive from regression-based causal mediation models using 1,000 bootstrap samples, assuming no exposure–mediator interaction and adjusted for the full covariate set.

For Lipodystrophy 1, a subtype reflecting adipose-tissue–driven insulin resistance, a broad set of lipid-related metabolites—including VLDL and LDL triglyceride fractions, HDL particle measures, PUFA-to-MUFA ratios, and apolipoprotein-related proteins—mediated a substantial proportion of the genetic effect on liver fat, pancreatic fat, and VAT mass. Several mediators accounted for 30–45% of the total effect, highlighting dysregulated lipoprotein remodeling and fatty-acid metabolism as central pathways linking genetic risk to ectopic fat accumulation. Similarly, mediation in Lipodystrophy 2 was dominated by triglyceride-rich lipoproteins and enzymes involved in lipid turnover. TG by PG and the hepatic enzyme ADH4 emerged as key mediators for liver fat, explaining 20–35% of the genetic effect. These results support distinct lipidomic architectures contributing to insulin resistance. (**Figure 6A**). In contrast, mediation for Beta Cell 1, a subtype representing genetically impaired insulin secretion, was minimal. Only GlycA, a marker of systemic inflammation, mediated a small portion (<20%) of the genetic association with VAT mass, and no mediator met criteria for pancreatic or liver fat. Lipodystrophy-related molecular features mediated 15–25% of the association between pPS and T2D risk, with prominent contributions from VLDL subclasses, lipid unsaturation indices, and lipid trafficking proteins (**Figure 6B**).

Finally, we assessed the stability of proteomic scores across three MESA examinations. Most scores showed substantial stability over time when tertiles were defined using Exam 1 cut points. The Beta Cell 2 proteomic score showed moderate tracking across Exams 1, 5, and 6, with most participants remaining in the same or an adjacent tertile and relatively few transitioning across extreme tertiles (**Supplementary Figure 7**).

## DISCUSSION

Type 2 diabetes is a biologically heterogeneous and multifactorial disease. Genetics has defined pathways of inherited susceptibility, but genetic risk is static and insufficient to explain the dynamic emergence and progression of disease. Circulating proteomic and metabolomic profiles, by contrast, capture the molecular consequences of genetic risk and its interaction with environmental and metabolic stress. Here, we demonstrate that integrating genomics with plasma multi-omics sharpens the biological interpretation of polygenic susceptibility to T2D. We identify distinct multi-omic signatures aligned with physiological genetic subtypes that capture molecular variation beyond genetic risk, track disease progression, and reveal subtype-specific clinical patterns that intensify in T2D. Notably, a β-cell–driven endotype defined by the Beta-Cell 2 multi-omic score showed marked stratification for insulin use, whereas the Obesity, Beta Cell 1, and Hyper Insulin scores were associated with cardiovascular complications, with effect sizes exceeding those observed for genetic risk scores alone. This highlights the potential of multi-omic profiling to advance risk stratification and clinically meaningful subtyping.

Our results demonstrate that the pPS are associated with distinct circulating proteomic and metabolomic profiles that mirror the fundamental biology of insulin resistance and insulin secretion. Insulin resistance pPS subtypes, particularly the Lipodystrophy 1 and Lipodystrophy 2 clusters, were characterized by perturbations in triglyceride-rich lipoproteins, altered fatty acid profiles, and activation of inflammatory pathways. For example, reduced levels of IGFBP2, a regulator of insulin sensitivity linked to obesity-related insulin resistance ^18,19^, were observed in the Lipodystrophy 1 cluster. In parallel, increased levels of TNFSF13, a pro-inflammatory mediator implicated in insulin resistance, and SMPD1^20,21^, a ceramide-generating enzyme known to disrupt insulin signaling, impair mitochondrial function, and promote inflammation, further support a mechanistic link to peripheral insulin insensitivity. Consistently, several proteins enriched within the insulin resistance pPS overlapped with established proteomic signatures of insulin sensitivity and insulin secretion^22^, including ACY1, LPE, and KITLG, providing external validation that these molecular patterns reflect bona fide insulin-resistant physiology. In contrast, pPS related to insulin secretion, represented by the Beta-Cell 1 and Beta-Cell 2 clusters, exhibited signatures enriched for proteins involved in beta-cell survival. Notably, lower circulating levels of CECAM1 protein, which support beta-cell survival and insulin secretory capacity ^23^, were observed within these clusters. Similarly, MET and LEP, key components of proteomic signatures associated with the acute insulin response to glucose (AIRg)^22^—a marker of beta-cell function—were also captured within the Beta-Cell 1 and Beta-Cell 2 pPS clusters. The clear molecular divergence between insulin resistance– and insulin secretion–driven pPS provides strong evidence that these scores capture biologically distinct and mechanistically coherent pathways underlying T2D heterogeneity. Our study builds on prior work focusing on an earlier iteration of five T2D genetic clusters and linear regression associations with proteomics (but not metabolomics)^24^, and replicates several of the notable proteomic associations from that study (e.g. reduced levels of IGFBP2 in Lipodystrophy cluster, reduced levels of CFC1 in Beta Cell cluster, increased FABP4 in Obesity cluster), in addition to offering novel associations.

While genetic scores capture fixed lifelong susceptibility, protein- and metabolite-based multi-omic scores reflect dynamic physiological states that are highly responsive to environmental and behavioral influences. ^25^ Consistent with this concept, prior studies have shown that proteomic risk scores achieve strong discrimination for common diseases, with C-indices exceeding 0.80 for multiple endpoints ^12^, and often match or outperform established clinical risk indicators. ^26^ In the context of T2D specifically, protein-based risk scores have been shown to outperform both polygenic risk scores and HbA1c ^11^, with additional studies demonstrating modest but meaningful improvements in prediction beyond traditional clinical and genetic factors. ^27^ In line with these observations, we found that pPS-based multi-omic scores tracked progressively across metabolic health states, from normoglycaemia through prediabetes to overt T2D, revealing subtype-specific molecular patterns that were amplified in individuals with established disease. Together, these findings highlight the value of multi-omic profiling as a dynamic and biologically proximal layer of risk that complements inherited genetic susceptibility. More broadly, they support a framework in which extending process-specific genetic scores through molecular profiling preserves mechanistic interpretability while sharpening associations with downstream molecular traits and clinically relevant outcomes ^28^.

Among the pPS multi-omic scores, the Beta-Cell 2 signature emerged as a strong predictor of insulin use and showed associations with cardiovascular outcomes, underscoring its potential value for risk stratification and disease monitoring. This finding highlights the ability to identify individuals in whom genetically anchored impairments in insulin secretory capacity led to progressive β-cell failure and eventual insulin dependence and cardiovascular damage. This pattern is biologically consistent with the central role of β-cell dysfunction in chronic hyperglycaemia, which promotes glucotoxicity, oxidative stress, and persistent low-grade inflammation.^29^

This study leverages the largest population-based resource with integrated genomic, detailed proteomic, and metabolomic data to define physiologically anchored multi-omic signatures of T2D using a robust LASSO-based framework with extensive validation, supported by causal inference analyses. External support in a multi-ancestry cohort strengthens the generalizability of these signatures. At the same time, longitudinal assessment demonstrates that key proteomic scores remain moderately stable over time, consistent with persistent underlying disease biology. The large effect sizes and complementary discrimination of multi-omic pPS indicate that these signatures capture biologically proximal disease states beyond inherited risk, highlighting their potential to inform patient stratification and mechanism-based prevention and therapeutic strategies. Future studies should extend this work across additional populations and molecular platforms, incorporate interventional omics, and determine the feasibility of implementing multi-omic pPS in clinical care. Additional efforts are warranted to evaluate the feasibility of implementing multi-omic risk stratification in clinical workflows and to explore the therapeutic potential of subtype-specific molecular pathways. Together, these steps will be critical for moving from multi-omic discovery toward precision prevention and treatment of T2D.

By integrating physiological polygenic pathway architecture with circulating proteomic and metabolomic profiles, we show that genetically anchored molecular signatures connect inherited susceptibility to downstream metabolic dysfunction and enable biologically interpretable subtyping of type 2 diabetes. This framework explains why individuals with comparable genetic risk can follow divergent clinical trajectories and develop distinct complication profiles. Ultimately, our findings demonstrate that multi-omic signatures can operationalize T2D heterogeneity, providing a foundation for mechanism-based risk stratification, prevention, and precision treatment.

## METHODS

### Study population

We analyzed data from the UK Biobank (UKBB), a large-scale, prospective cohort study that recruited ∼500,000 participants between 2006 and 2010 through the United Kingdom’s National Health Service. At enrollment, participants were 40–69 years of age and underwent extensive phenotyping through questionnaires, physical examinations, and the collection of biological samples, as described in detail elsewhere.^30^ The UK Biobank study was approved by the North West Multi-Centre Research Ethics Committee, and all participants provided written informed consent. The full study protocol is publicly available (https://www.ukbiobank.ac.uk). Analysis of the UK Biobank was conducted under application # 27892.

### Genotyping and Quality Control

All UKBB participants underwent genome-wide genotyping. Details of genotyping arrays, imputation procedures, and quality control protocols for the genetic dataset have been reported previously.^31^

### Metabolomic Data

Baseline plasma samples from all 502,000 UK Biobank participants were profiled by Nightingale Health Plc. using their high-throughput nuclear magnetic resonance (NMR) metabolomics platform. This platform quantifies a broad range of metabolic traits, including lipids, fatty acids, amino acids, glycolysis-related metabolites, and inflammatory markers. In total, 249 biomarkers were measured. Detailed descriptions of the Nightingale Health NMR biomarker platform in the UK Biobank have been published previously.^32^ Derived metabolite variables expressed as percentages were excluded from all analyses. Glucose was also excluded, given its direct role in the clinical diagnosis of T2D and the potential for circularity in exposure and outcome definitions. The total number of metabolites considered in downstream analysis was 191 (**Supp Table 6**).

### Proteomic Data

Proteomic profiling was available for 53,021 UK Biobank participants and was performed using Olink’s Proximity Extension Assay (PEA) technology. In this assay, pairs of antibodies labeled with complementary oligonucleotides (proximity probes) selectively bind to their target proteins in plasma. In total, 2,923 unique proteins were measured across panels targeting inflammation (INF, INF II), oncology (ONC, ONC II), cardiometabolic (CAR, CAR II), and neurological (NEU, NEU II) pathways. Details of sample handling, laboratory protocols, and the Olink proteomic platform in the UK Biobank have been described elsewhere.^33^ Raw counts were transformed into Normalized Protein eXpression (NPX) values, reported on an arbitrary log2 scale to facilitate relative quantification across samples. The final list of proteins can be found in **Supplementary Table 7**.

### Generation of genetic partitioned polygenic scores (pPS)

We constructed 12 partitioned polygenic risk scores (pPS), each capturing genetic susceptibility to T2D through distinct pathophysiological pathways, using previously published weights^4^. In addition, we selected a global T2D polygenic risk score (PRS) derived from GWAS summary statistics excluding UK Biobank participants, thereby minimizing the risk of overfitting in our analyses.^34^

Both the global and pathway-specific pPS were calculated as weighted sums of T2D-associated genetic variants. We primarily used genotyped variants directly; when these were unavailable, imputed variants were incorporated. For variants not available, we selected proxy variants with high linkage disequilibrium (r² > 0.8). The computational pipeline used to generate these scores is publicly available (https://github.com/manning-lab/pprs-pipeline).

### Generation of multi-omic signatures

To identify multi-omic signatures representing the polygenic physiologic score (pPS), we applied least absolute shrinkage and selection operator (LASSO) regression across three analytic layers: metabolites alone, proteins alone, and an integrated metabolite–protein dataset (primary analysis). Plasma metabolomic and proteomic measurements obtained at the first assessment were used for model derivation.

The analytic sample for training the LASSO regression was restricted to participants free of major chronic disease. Specifically, we excluded individuals who (i) reported a history of coronary heart disease or stroke, type 1 or type 2 diabetes (UK Biobank fields 6150 and 2443, instance 0), or a cancer diagnosis with a recorded date prior to baseline (field 40005); or (ii) had baseline HbA1c >6.5% or reported insulin use at baseline (field 6177 for men and field 6153 for women, instance 0). Within the training set, we fit a LASSO model (α = 1) and optimized the penalty parameter λ using a nested cross-validation framework. For each of the 10 outer folds, an inner 10-fold cross-validation was performed to identify the optimal λ based on the minimum cross-validation error criterion. The median of these λ min values was selected as the final penalty parameter and used to refit the LASSO model on the full training set. In the test set, multi-omic scores were calculated as weighted sums of the selected biomarkers, with weights corresponding to the LASSO-derived coefficients. To ensure adequate genetic information content, score analyses were restricted to pPS clusters comprising more than 20 SNPs (Beta Cell 1 and 2, Obesity, Hyper Insulin, Lipodystrophy 1 and 2).

The testing set included participants across the full glycemic spectrum, enabling model development and evaluation of disease-stage performance. Model significance was later evaluated using permutation testing (see Statistical Analysis).

### Pathway Enrichment Analysis

We conducted pathway enrichment analysis of the selected pPS multi-omic signatures using the Reactome pathway database, implemented through the Enrichr tool. ^35^ To capture biologically meaningful patterns, we performed enrichment separately for biomarkers with positive and negative LASSO weights, thereby distinguishing pathways associated with up-versus down-weighted features in the multi-omic signatures.

### Outcome definition

Prevalent T2D at baseline was defined as a recorded diagnosis of T2D (ICD-10 codes E11.0–E11.9) in hospital inpatient records (UK Biobank field 41270) before or at the baseline assessment visit. Non-T2D was defined as no diagnosis of T2D at baseline or follow-up and no reported use of diabetes medication throughout follow-up.

Prediabetes was defined using fasting plasma glucose and HbA1c at visit 1, in accordance with American Diabetes Association criteria^36^. Participants were classified as having prediabetes if glucose levels were 5.6–6.9 mmol/L or HbA1c 39–47 mmol/mol, had no diagnosis of diabetes, and were not on diabetes medications; those with glucose <5.6 mmol/L and HbA1c <39 mmol/mol were classified as normoglycemic. Individuals with baseline type 2 diabetes or missing/discordant values were not assigned to the prediabetes category.

Insulin use was ascertained from UKBB medication records using insulin-specific medication codes at the initial assessment (field IDs 6153 and 6177). Participants were classified as insulin users if they reported current use of any insulin preparation. In addition, a cumulative ever-insulin variable was derived to indicate insulin use reported at any visit.

Incident cardiovascular disease was defined as a myocardial infarction diagnosis recorded after the assessment visit, based on self-report in UK Biobank Field 20002 and hospital records using ICD-10 codes I21–I25 and ICD-9 codes 410–412 and 429.79.

### Assessment of Clinical Biomarkers

We selected a panel of biomarkers representing key domains of metabolic health to characterize the phenotypic profile associated with the multi-omic pPS. Clinical measurements obtained at the baseline visit included LDL cholesterol, HDL cholesterol, triglycerides, alanine aminotransferase, aspartate aminotransferase, gamma-glutamyltransferase, C-reactive protein, glucose, total cholesterol, BMI, HbA1c, waist circumference, hip circumference, and waist-hip ratio. Kidney function was estimated using the CKD-EPI eGFR^37^ equation based on serum creatinine. Insulin resistance was approximated using the triglyceride-to-HDL cholesterol ratio, calculated from triglyceride and HDL cholesterol measurements.^38^ Body composition measures, including visceral adipose tissue, trunk fat mass, and total fat mass, were derived from DXA scans conducted at Visit 2, and liver fat content was quantified from MRI scans at Visit 2.

### Statistical Analysis

Individuals with >10% missingness in either metabolomic or proteomic data were excluded. For metabolites, missing values were imputed using half of the minimum observed concentration, followed by inverse normal transformation for normalization. For proteins, missing values were imputed using a 10 nearest-neighbor approach and similarly normalized with inverse normal transformation. Clinical biomarkers were transformed using an inverse rank normal transformation to approximate normality.

To assess whether the predictive performance of each multi-omic pPS model exceeded that expected by chance, we implemented permutation testing. The outcome label (pPS category) was randomly shuffled in the training set, the model was refit using the same penalty parameter (λ) selected from the original cross-validation, and the prediction was evaluated in the test set.

This procedure was repeated 1,000 times to generate a null distribution of model performance, against which the observed performance was compared to compute permutation-based p-values.

For each multi-omic pPS, we quantified overlap between cluster-specific multi-omic features and the global T2D feature set. Fold enrichment was calculated as the observed overlap relative to the expected overlap under random feature assignment. Statistical significance was assessed using a hypergeometric test with false discovery rate (FDR) correction for multiple testing.

Correlations between the genetic and multi-omic pPS scores, as well as within multi-omics pPS were quantified using Pearson correlation coefficients. To compare their predictive contributions, we evaluated type 2 diabetes prediction across three models: a genetic pPS–only model, a multi-omic pPS–only model, and a combined model including both scores. Incremental discrimination was assessed by changes in AUC, and statistical significance of AUC differences was tested using the DeLong method.

To examine whether the two scores capture independent components of diabetes risk, we fit joint logistic regression models including both pPS scores simultaneously. Multiplicative interactions were assessed by adding a product term between the genetic and multi-omic scores.

To compare the extent to which omics and genetic scores captured subtype-defining endotypes, we tested their associations with clinical traits using linear regression in individuals without T2D and in those with T2D. Exposures were standardized, outcomes inverse-normal transformed, and models adjusted for age, sex, and 10 genetic principal components; glucose analyses were restricted to participants fasting for more than 6 h, and false discovery rate was controlled using the Benjamini–Hochberg method within each analysis group and outcome. We also examined variation in multi-omic pPS across metabolic disease stages (no disease, prediabetes, and T2D) using linear trend tests and assessed their associations with key metabolic traits overall and in stratified analyses to determine whether they recapitulated the clinical features represented by their genetic counterparts. Effect modification by disease status was tested using a pPS × disease stage interaction term.

We modelled prevalent T2D for each score separately using logistic regression adjusted for age, sex, 10 genetic principal components, and fasting hours. Individuals were grouped into percentiles of each score, and the mean fitted probability of prevalent T2D was calculated within each percentile.

To evaluate associations between subtype-specific scores and baseline insulin use, we created binary indicators identifying individuals in the top 10% of each score distribution and compared them with the remaining 90% of participants. Separate logistic regression models were then fit for each score. To maintain a consistent analytic sample across scores, all models were restricted to individuals with available multi-omic score data. Models were adjusted for age, sex, and the first 10 genetic principal components. Odds ratios and 95% confidence intervals were estimated for insulin use in the top 10% versus the remainder of the distribution.

The cumulative proportion of insulin use was assessed using age at insulin initiation, which was calculated from the earliest recorded insulin date relative to birth date, with birth date approximated as the 15th day of the reported birth month and year, and another analysis relative to the years since T2D diagnosis. Among individuals without insulin use, follow-up was censored at the recorded censoring date, and age at censoring was calculated analogously. To assess progression to insulin use, each score was categorized into tertiles, and participants were assigned to mutually exclusive groups based on unique top-tertile membership, defined as belonging to the highest tertile of one score only. Individuals in the highest tertile of more than one score were excluded. Differences across groups were evaluated using a global log-rank test. Adjusted cumulative incidence curves were then estimated using Cox proportional hazards models with age as the time scale, adjusting for sex, years living with T2D (for the analysis relative to birth date), and the first 10 genetic principal components. Model-based cumulative probabilities were obtained as 1 minus the estimated survival probability.

Finally, we used logistic regression models to evaluate the association of the scores with incident cardiovascular disease, adjusting for age, sex, 10 genetic principal components, statin use, and smoking quantity and frequency. Statistical significance was set at *P < 0.004* after correcting for multiple testing across six scores and two T2D-related complications.

### Mediation analysis

To investigate potential causal pathways linking the genetic pPS and their molecular signatures with type 2 diabetes and regional adiposity traits, we conducted mediation analyses using the cmest function from the CMAverse package with statistical inference derived from 1,000 bootstrap samples. ^39^ Models were specified under the assumption of no exposure–mediator interaction. Potential mediators were selected based on their significant associations with both the exposure and the outcome, enabling estimation of the extent to which multi-omic signatures transmit the effects of genetic predisposition on metabolic outcomes. Genetic pPS were modeled as exposures, with their corresponding multi-omic pPS features included as mediators. Liver fat, visceral adipose tissue, trunk fat mass, and total fat mass were grouped as regional adiposity traits, representing fat accumulation across physiologically distinct depots with differential metabolic relevance. Adiposity traits measured at Visit 2 and incident T2D were used as outcomes to ensure temporal ordering consistent with causal mediation assumptions. All models were adjusted for the full covariate set. A mediation effect was considered statistically significant when the proportion mediated exceeded 10% with a corresponding *P* <1x 10□^3^. All analyses were conducted in R (version 4.3).

### Replication Cohort and Analysis

Key findings were evaluated in the Multi-Ethnic Study of Atherosclerosis (MESA), a population-based multi-ethnic cohort with extensive phenotypic and molecular Detailed study design information is provided in the Supplementary Methods and has been described previously^40^. The Institutional Review Boards approved the study at all participating institutions, and all participants provided written informed consent. Plasma proteomic profiling was performed using the Olink Explore 3072 platform, matching the proteomic panel used in UK Biobank. We constructed two sets of scores in MESA using weights from the UK Biobank testing subset: protein-only scores and modified multi-omic scores, in which metabolite weights were set to zero because an equivalent metabolomic panel was not available in MESA. Protein levels were standardized using the means and standard deviations from the UK Biobank testing subset. Incident T2D was defined as the first occurrence after proteomic measurement of fasting glucose >7.0 mmol/L after at least 8 hours of fasting, initiation of diabetes treatment, hemoglobin A1c >6.5%, physician diagnosis of diabetes, or self-reported diabetes. Insulin use was defined based on self-report at Exam 1 (baseline), Exam 5 (approximately 10 years after baseline), and Exam 6 (approximately 15 years after baseline). Clinical characteristics are shown in **Supplementary Table 8**. Cox proportional hazards models were used to evaluate associations between baseline ‘omic scores and incident T2D. Models were adjusted for age, sex, study site, and the first 10 genetic principal components. Logistic regression was used to assess associations between baseline ‘omic scores and insulin use, with adjustment for the same covariates.

## Data Availability

All data produced in the present work are contained in the manuscript

## CODE AND DATA AVAILABILITY

https://github.com/manning-lab/pprs-pipeline

All data produced in the present work are contained in the manuscript

## ACKNOWLEDGMENTS

The authors are grateful to UK Biobank for access to data to undertake this study (Project #30418). This work uses data provided by patients and collected by the NHS as part of their care and support. Nightingale Health Plc is acknowledged for early access to the UK Biobank NMR biomarker data and discussions regarding sources of experimental variation.

Special acknowledgements to the Trans-Omics in Precision Medicine (TOPMed) program, supported by the National Heart, Lung and Blood Institute (NHLBI). NHLBI TOPMed: Multi-Ethnic Study of Atherosclerosis (MESA)” (phs001416.v1.p1) was performed at the Broad Institute of MIT and Harvard (3U54HG003067-13S1). Centralized read mapping and genotype calling, along with variant quality metrics and filtering were provided by the TOPMed Informatics Research Center (3R01HL-117626-02S1, contract HHSN268201800002I) (Broad RNA Seq, Proteomics HHSN268201600034I, UW RNA Seq HHSN268201600032I, USC DNA Methylation HHSN268201600034I, Broad Metabolomics HHSN268201600038I). Phenotype harmonization, data management, sample-identity QC, and general study coordination, were provided by the TOPMed Data Coordinating Center (3R01HL-120393; U01HL-120393; contract HHSN268180001I). MESA and the MESA SHARe projects are conducted and supported by the National Heart, Lung, and Blood Institute (NHLBI) in collaboration with MESA investigators. Support for MESA is provided by contracts 75N92025D00022, 75N92020D00001, HHSN2682015000031/HSN26800004, N01-HC-95159, 75N92025D00026, 75N92020D00005, N01-HC-95160, 75N92020D00002, N01-HC-95161, 75N92025D00024, 75N92020D00003, N01-HC-95162, 75N92025D00027, 75N92020D00006, N01-HC-95163, 75N92025D00025, 75N92020D00004, N01-HC-95164, 75N92025D00028, 75N92020D00007, N01-HC-95165, N01-HC-95166, N01-HC-95167, N01-HC-95168, N01-HC-95169, UL1-TR-000040, UL1-TR-001079, UL1-TR-001420, UL1TR001881, R01DK081572, R01HL133870, and R01HL105756. Funding for SHARe genotyping was provided by NHLBI Contract N02-HL-64278. The authors thank the MESA participants and the MESA investigators and staff for their valuable contributions. A full list of participating MESA investigators and institutions can be found at http://www.mesa-nhlbi.org.

