## Supplementary Methods for "Multi-omic signatures of genetic mechanisms inform on type 2 diabetes biology and patient heterogeneity"

The Multi-Ethnic Study of Atherosclerosis (MESA) is a study of the characteristics of subclinical cardiovascular disease and the risk factors that predict progression to clinically overt cardiovascular disease or progression of the subclinical disease ^41^. MESA consists of a diverse, community-based sample of an initial 6,814 men and women aged 45-84 years without known cardiovascular disease at baseline. Thirty-eight percent of the recruited participants were White, 28 percent African American, 22 percent Hispanic, and 12 percent of Chinese descent. Participants were recruited from six field centers across the United States: Baltimore City and Baltimore County, Maryland; Chicago, Illinois; Forsyth County, North Carolina; Los Angeles County, California; New York, New York; and St. Paul, Minnesota. The first examination took place over two years, from July 2000 to July 2002, and has been followed by additional examinations.

Participants are being followed for identification and characterization of cardiovascular disease events, including acute myocardial infarction and other forms of coronary heart disease (CHD), stroke, and heart failure; for cardiovascular disease interventions; and for mortality. Follow-up telephone interviews with participants or their proxies are attempted at least annually to identify new hospitalizations and diagnoses. MESA staff request information on hospital admissions for any reason, outpatient CVD diagnoses, and death. Death certificate records are obtained from state vital statistics or the National Death Index. MESA staff record International Classification of Disease (ICD) diagnosis codes for all hospitalizations and request medical records for those with ICD codes related to CVD. MESA staff also request outpatient records for potential CVD events. However, not all hospitalizations and/or CVD event records were fully documented.

The study was approved by the Institutional review boards at all participating institutions, and all participants gave written informed consent. In addition, informed consent was obtained for extensive data sharing (dbGaP) and genetic/omic studies, including candidate genes (NHLBI CARe), genome-wide scans (NHLBI SHARe), exome sequencing (NHLBI ESP) and, most recently, the NHLBI TOPMed program.
