## Supplementary Figures for "Multi-omic signatures of genetic mechanisms inform on type 2 diabetes biology and patient heterogeneity"

### Slide 1
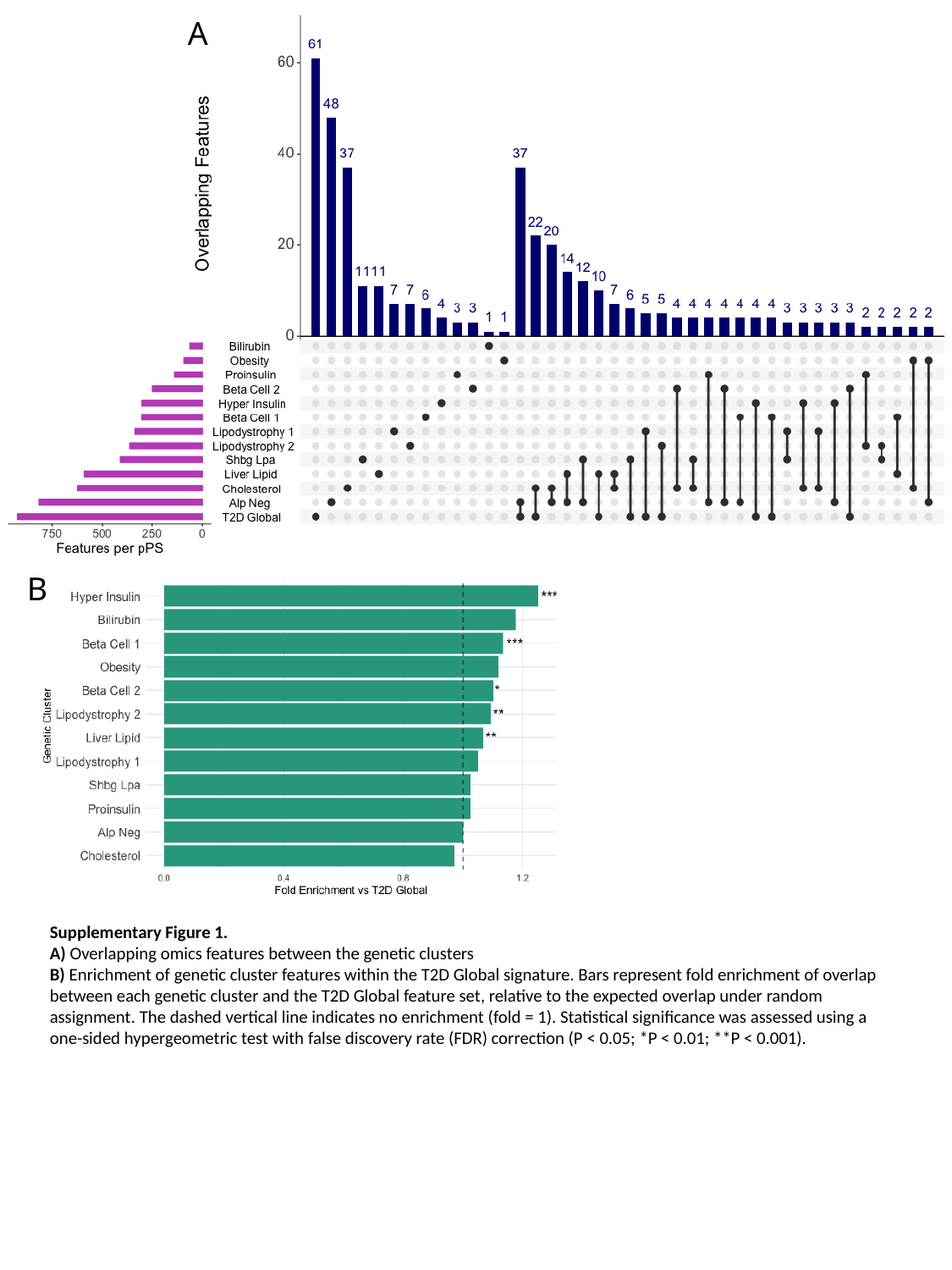

A
B
Supplementary Figure 1.
A) Overlapping omics features between the genetic clusters
B) Enrichment of genetic cluster features within the T2D Global signature. Bars represent fold enrichment of overlap between each genetic cluster and the T2D Global feature set, relative to the expected overlap under random assignment. The dashed vertical line indicates no enrichment (fold = 1). Statistical significance was assessed using a one-sided hypergeometric test with false discovery rate (FDR) correction (P < 0.05; *P < 0.01; **P < 0.001).

### Slide 2
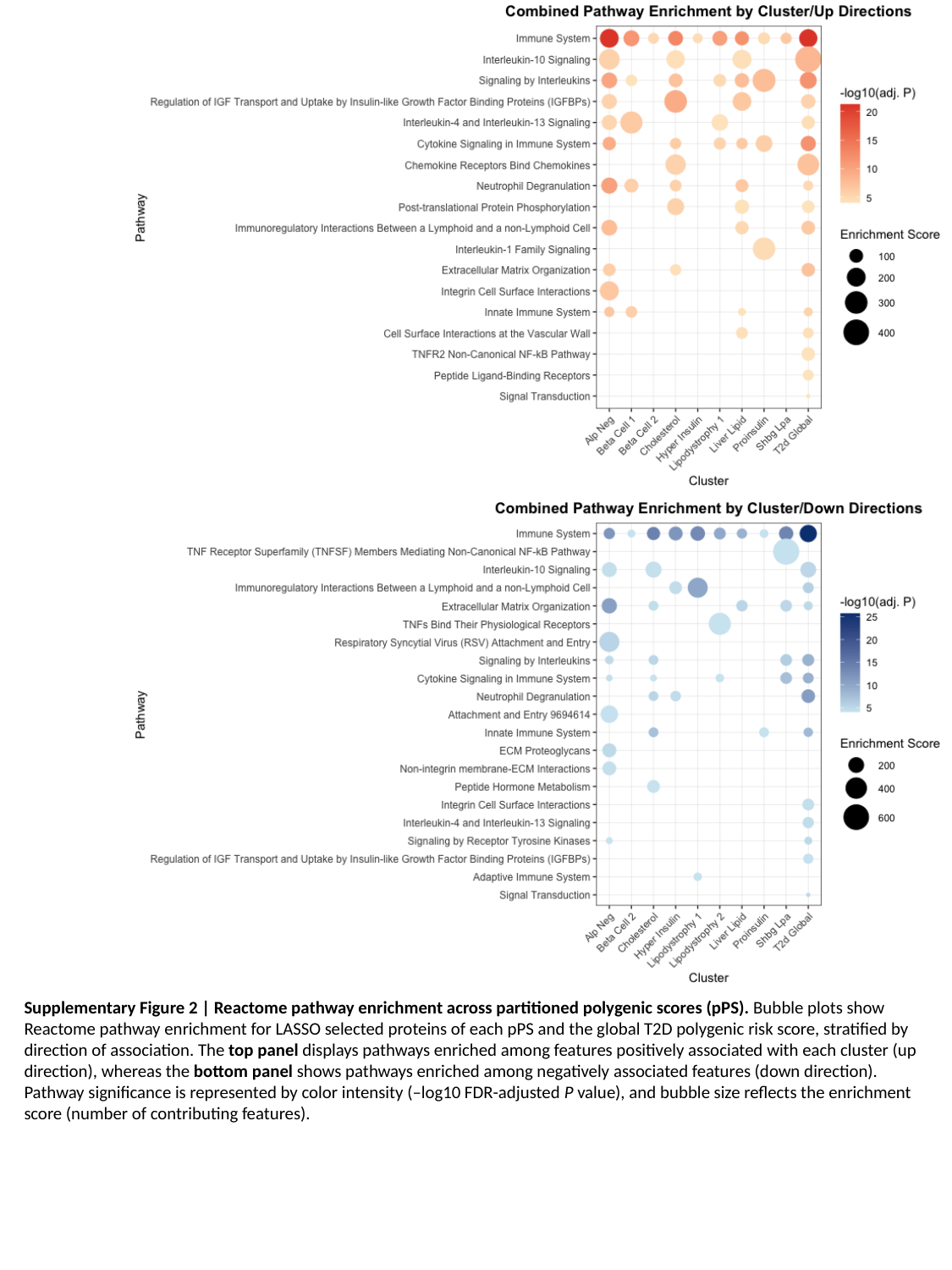

Supplementary Figure 2 | Reactome pathway enrichment across partitioned polygenic scores (pPS). Bubble plots show Reactome pathway enrichment for LASSO selected proteins of each pPS and the global T2D polygenic risk score, stratified by direction of association. The top panel displays pathways enriched among features positively associated with each cluster (up direction), whereas the bottom panel shows pathways enriched among negatively associated features (down direction). Pathway significance is represented by color intensity (–log10 FDR-adjusted P value), and bubble size reflects the enrichment score (number of contributing features).

### Slide 3
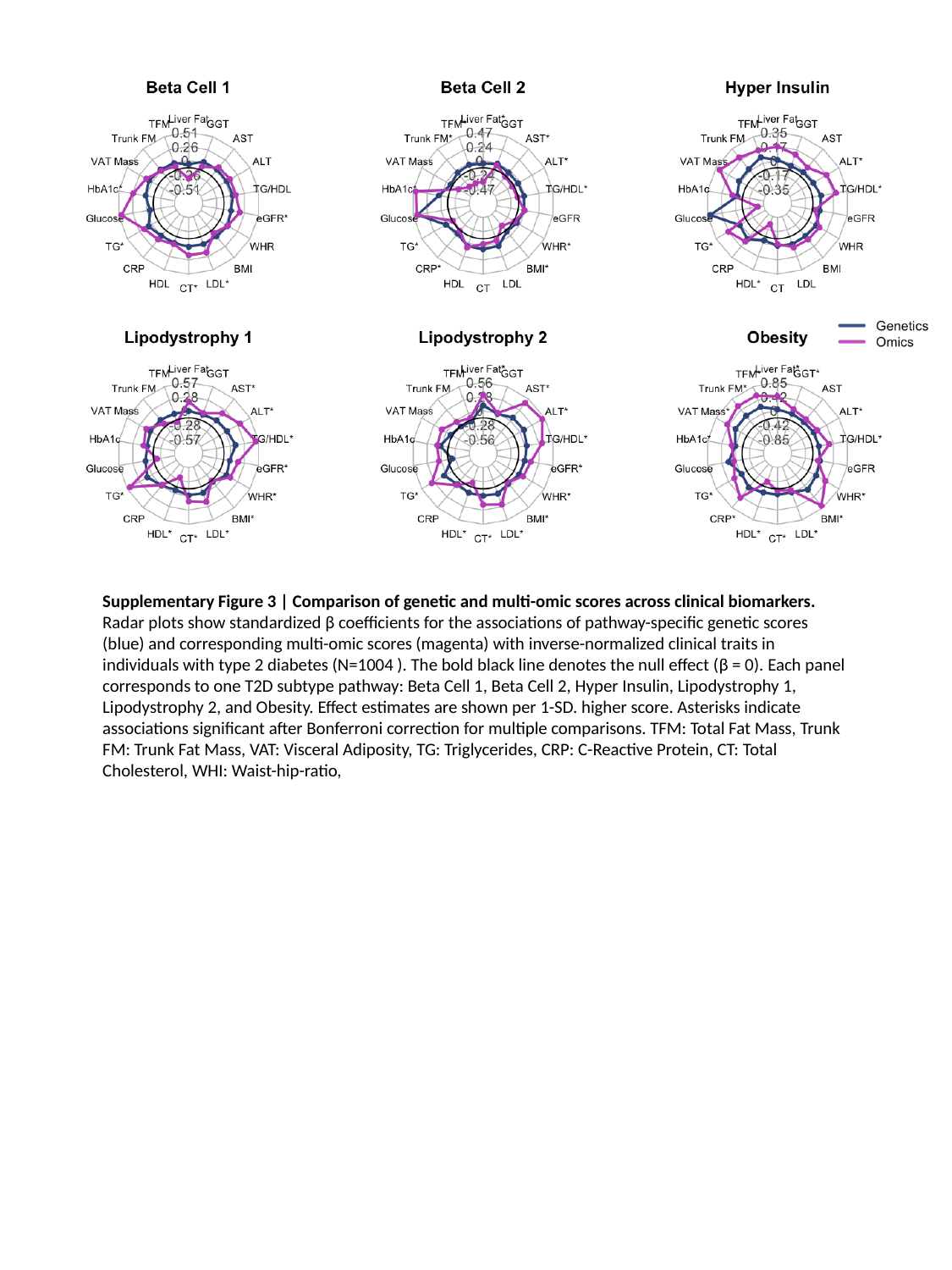

Supplementary Figure 3 | Comparison of genetic and multi-omic scores across clinical biomarkers. Radar plots show standardized β coefficients for the associations of pathway-specific genetic scores (blue) and corresponding multi-omic scores (magenta) with inverse-normalized clinical traits in individuals with type 2 diabetes (N=1004 ). The bold black line denotes the null effect (β = 0). Each panel corresponds to one T2D subtype pathway: Beta Cell 1, Beta Cell 2, Hyper Insulin, Lipodystrophy 1, Lipodystrophy 2, and Obesity. Effect estimates are shown per 1-SD. higher score. Asterisks indicate associations significant after Bonferroni correction for multiple comparisons. TFM: Total Fat Mass, Trunk FM: Trunk Fat Mass, VAT: Visceral Adiposity, TG: Triglycerides, CRP: C-Reactive Protein, CT: Total Cholesterol, WHI: Waist-hip-ratio,

### Slide 4
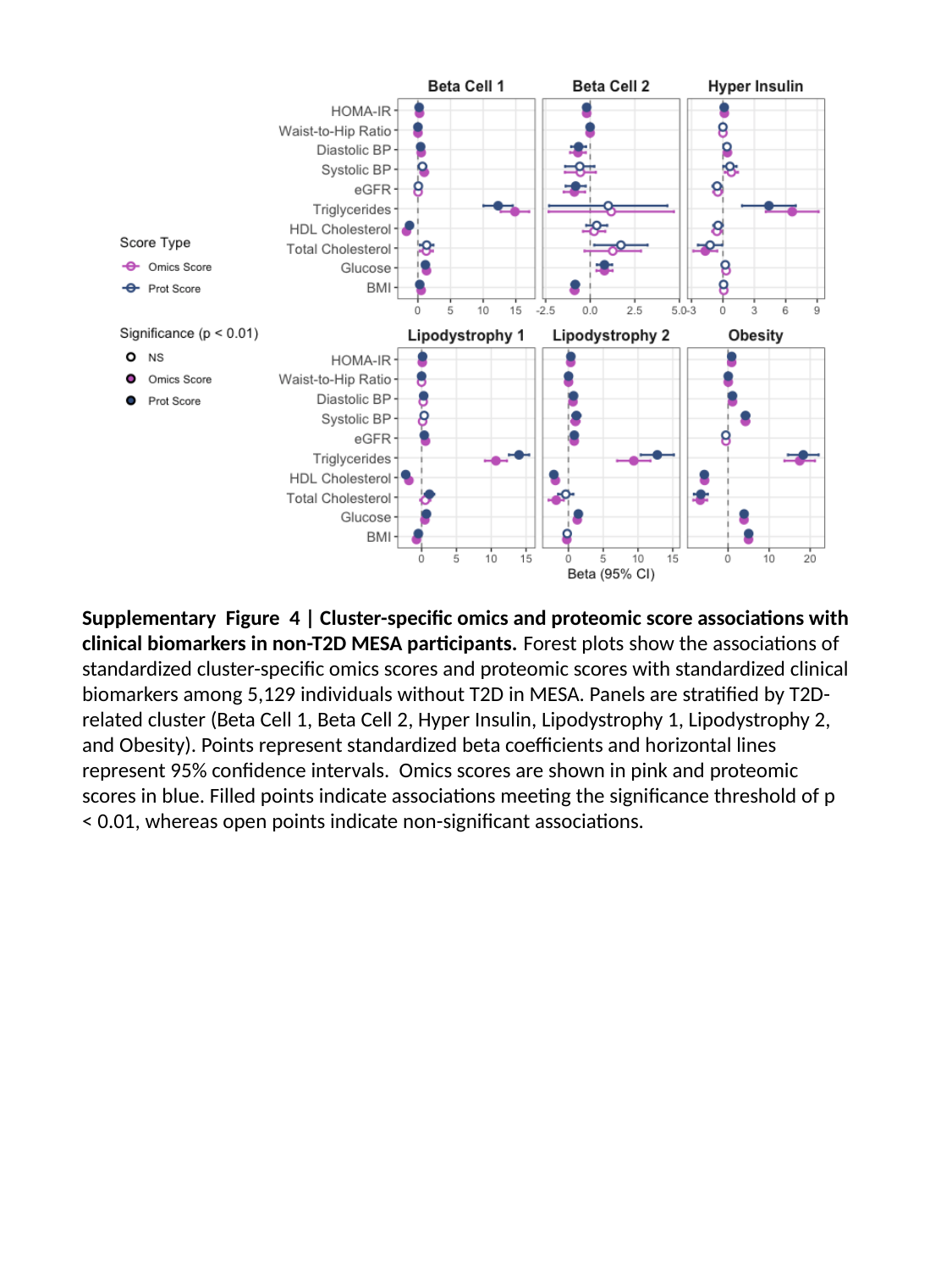

Supplementary Figure 4 | Cluster-specific omics and proteomic score associations with clinical biomarkers in non-T2D MESA participants. Forest plots show the associations of standardized cluster-specific omics scores and proteomic scores with standardized clinical biomarkers among 5,129 individuals without T2D in MESA. Panels are stratified by T2D-related cluster (Beta Cell 1, Beta Cell 2, Hyper Insulin, Lipodystrophy 1, Lipodystrophy 2, and Obesity). Points represent standardized beta coefficients and horizontal lines represent 95% confidence intervals. Omics scores are shown in pink and proteomic scores in blue. Filled points indicate associations meeting the significance threshold of p < 0.01, whereas open points indicate non-significant associations.

### Slide 5
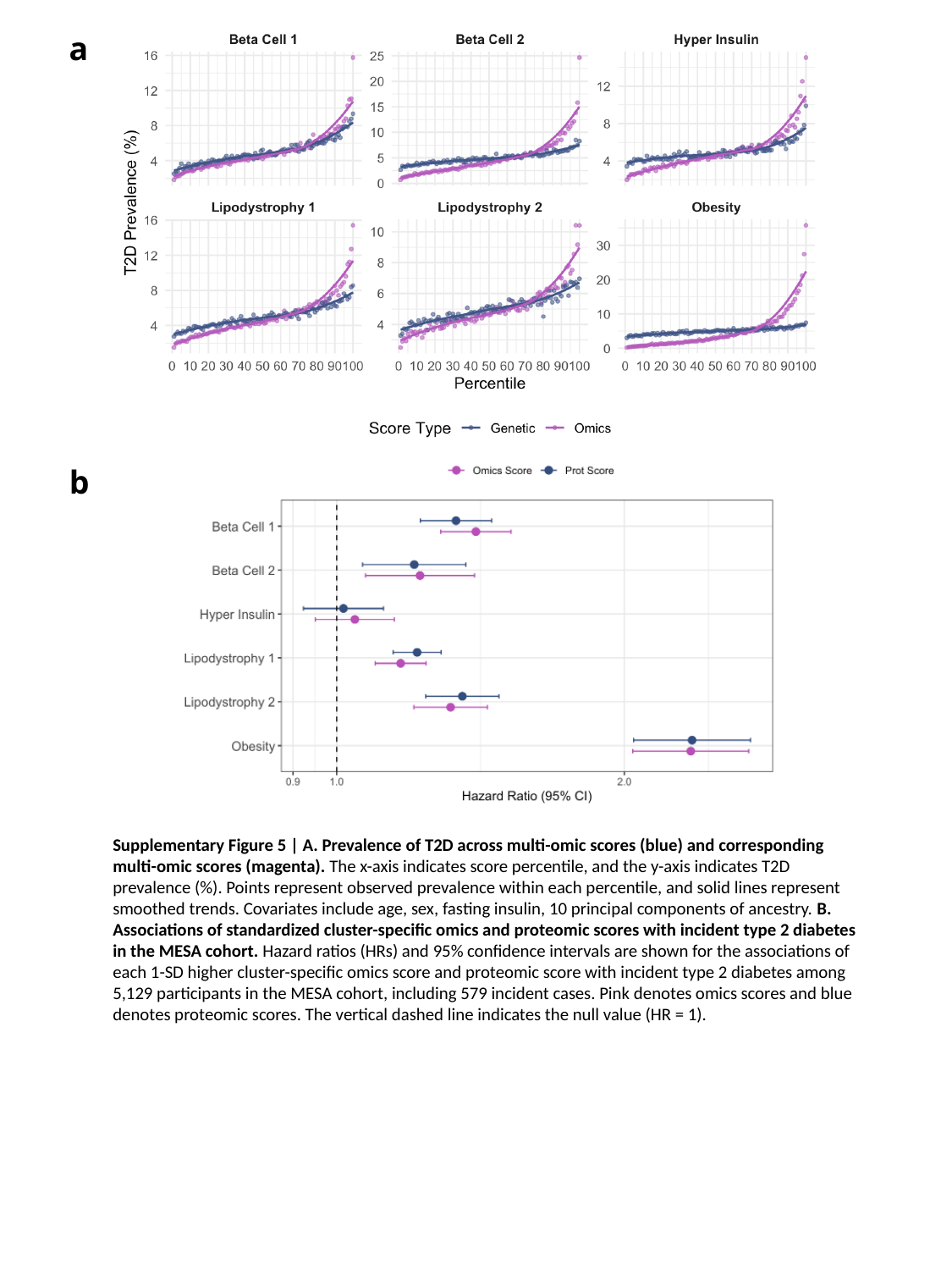

a
b
Supplementary Figure 5 | A. Prevalence of T2D across multi-omic scores (blue) and corresponding multi-omic scores (magenta). The x-axis indicates score percentile, and the y-axis indicates T2D prevalence (%). Points represent observed prevalence within each percentile, and solid lines represent smoothed trends. Covariates include age, sex, fasting insulin, 10 principal components of ancestry. B. Associations of standardized cluster-specific omics and proteomic scores with incident type 2 diabetes in the MESA cohort. Hazard ratios (HRs) and 95% confidence intervals are shown for the associations of each 1-SD higher cluster-specific omics score and proteomic score with incident type 2 diabetes among 5,129 participants in the MESA cohort, including 579 incident cases. Pink denotes omics scores and blue denotes proteomic scores. The vertical dashed line indicates the null value (HR = 1).

### Slide 6
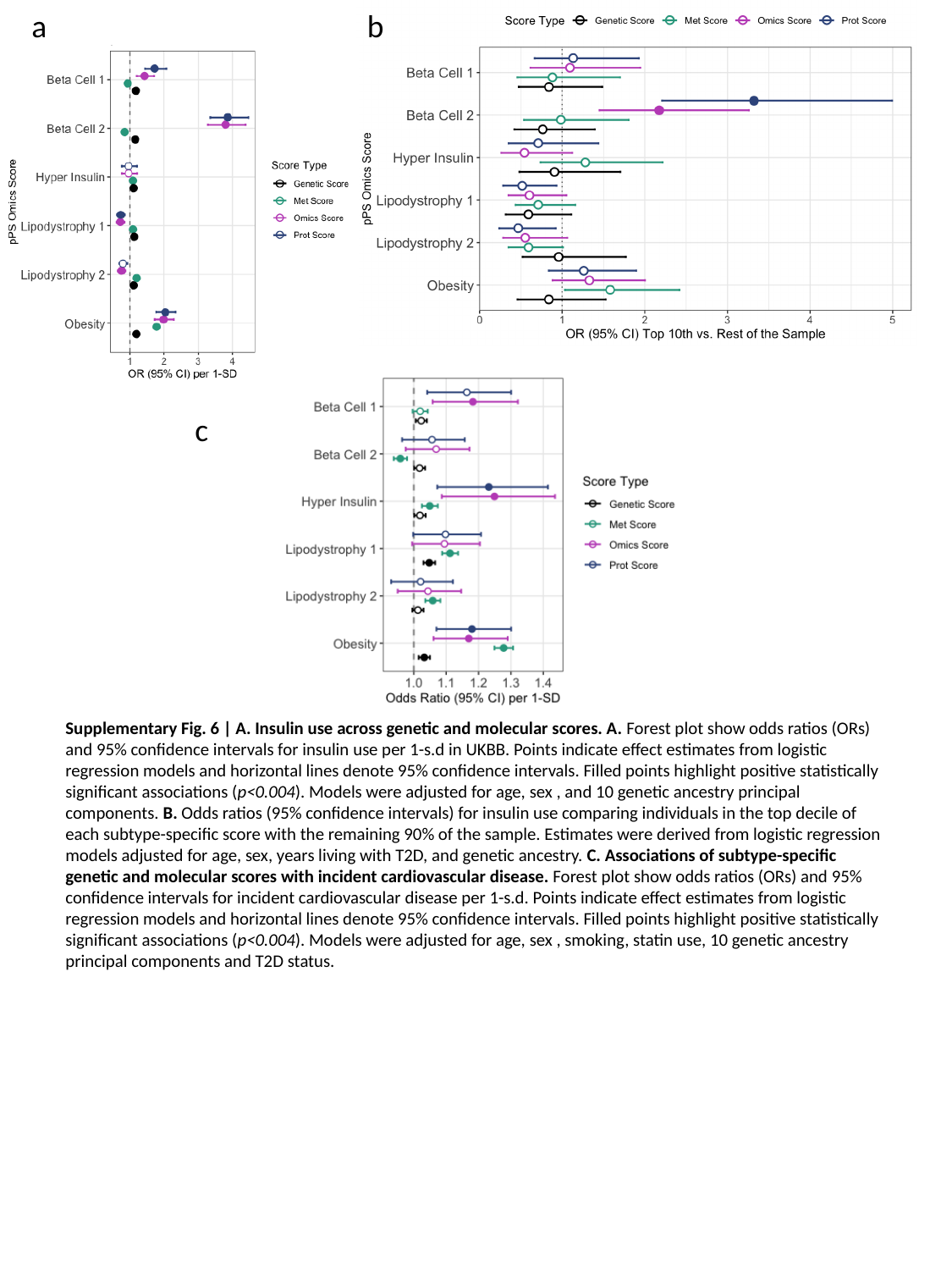

a
b
c
Supplementary Fig. 6 | A. Insulin use across genetic and molecular scores. A. Forest plot show odds ratios (ORs) and 95% confidence intervals for insulin use per 1-s.d in UKBB. Points indicate effect estimates from logistic regression models and horizontal lines denote 95% confidence intervals. Filled points highlight positive statistically significant associations (p<0.004). Models were adjusted for age, sex , and 10 genetic ancestry principal components. B. Odds ratios (95% confidence intervals) for insulin use comparing individuals in the top decile of each subtype-specific score with the remaining 90% of the sample. Estimates were derived from logistic regression models adjusted for age, sex, years living with T2D, and genetic ancestry. C. Associations of subtype-specific genetic and molecular scores with incident cardiovascular disease. Forest plot show odds ratios (ORs) and 95% confidence intervals for incident cardiovascular disease per 1-s.d. Points indicate effect estimates from logistic regression models and horizontal lines denote 95% confidence intervals. Filled points highlight positive statistically significant associations (p<0.004). Models were adjusted for age, sex , smoking, statin use, 10 genetic ancestry principal components and T2D status.

### Slide 7
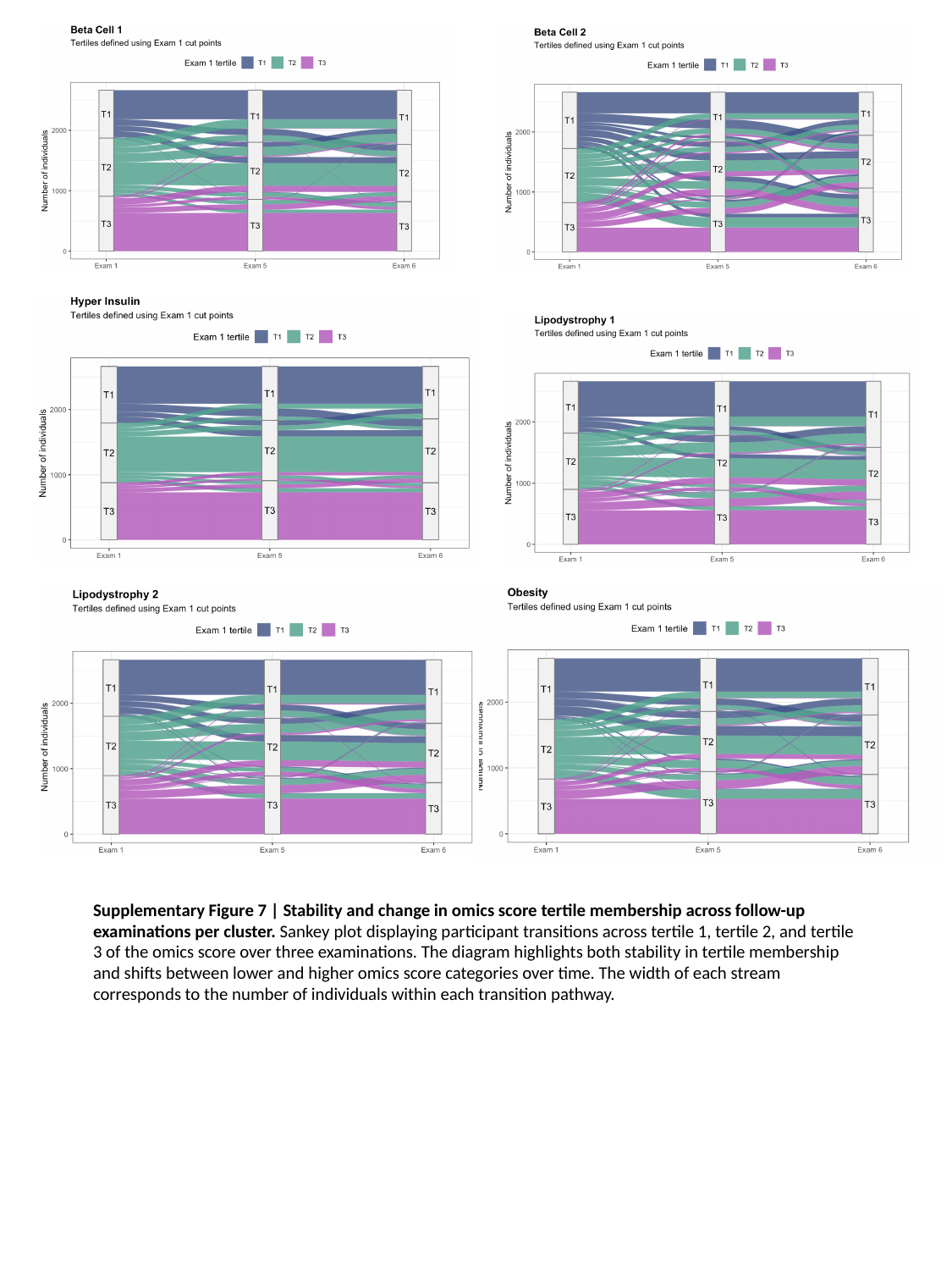

Supplementary Figure 7 | Stability and change in omics score tertile membership across follow-up examinations per cluster. Sankey plot displaying participant transitions across tertile 1, tertile 2, and tertile 3 of the omics score over three examinations. The diagram highlights both stability in tertile membership and shifts between lower and higher omics score categories over time. The width of each stream corresponds to the number of individuals within each transition pathway.
